# A Multivariable Plasma Extracellular Vesicle Surface Profile Associated with Post-COVID-19 Syndrome

**DOI:** 10.64898/2026.08.27.26361498

**Authors:** Deborah K. Erhart, Hanna Reßin, Luisa T. Balz, Shreyans Chatterjee, Dorothée Lulé, Susanne Müller, Jan Lewerenz, Jan Münch, Hayrettin Tumani, Rüdiger M. Groß

## Abstract

Post-COVID-19 syndrome (PCS) is characterized by fatigue, neurological impairment and systemic symptoms. This heterogeneity of symptoms hinders biomarker development. Here, we profiled extracellular-vesicle (EV) surface markers in plasma and CSF from 61 participants with PCS (COVID^post^), 80 recovered controls (COVID^reco^), and 10 participants with non-SARS-CoV-2 post-viral syndromes. EVs were analysed by bead-based multiplex flow cytometry using tetraspanin-directed (TSPN) and phosphatidylserine-directed lactadherin (PS) detection. Amongst 37 targets covering tetraspanins and vasculature-, immunity- and stemness-associated markers, none met a 1% false-discovery-rate threshold. However, L1-regularized logistic regression under fully nested 5×5 cross-validation identified a distributed plasma EV profile, with mean out-of-fold areas under the receiver operating characteristic curve (AUCs) of 0.788 (95% CI 0.715–0.852) for TSPN and 0.716 (95% CI 0.636–0.792) for PS detection. Across the pooled COVID^post^ and COVID^reco^ population, EV classification scores covaried with clinical group differences, but did not track clinical severity within either cohort. These PCS-EV classification scores decreased at one-year follow-up in COVID^post^ participants. Our findings identify an internally cross-validated multivariable EV surface profile associated with COVID^post^ versus COVID^reco^ status and support independent validation and exploration of EV-based biomarkers in post-viral fatigue syndromes.

## Introduction

The COVID-19 pandemic, caused by severe acute respiratory syndrome coronavirus 2 (SARS-CoV-2), has resulted in over 700 million confirmed infections worldwide. A substantial proportion of patients experience persistent or newly arising symptoms long after viral clearance, a condition termed post-COVID-19 syndrome (PCS), which usually occurs three months after the onset of SARS-CoV-2 infection, with symptoms lasting for at least two months and no alternative explanation^1^. PCS encompasses over 200 reported symptoms affecting multiple organ systems, with fatigue, cognitive impairment, post-exertional malaise and autonomic dysfunction among the most disabling manifestations^2^. High-dimensional analyses have confirmed excess risk across pulmonary, cardiovascular, neurological, metabolic, and gastrointestinal domains, with risk graded by acute disease severity^3^. Conservative estimates suggest that at least 10% of infected individuals develop persistent symptoms, translating to tens of millions of affected people globally^2,3^. The resulting burden on patients and healthcare systems is enormous, with affected individuals frequently reporting reduced or completely abrogated work capacity and diminished quality of life^2^. At present, diagnosis remains entirely clinical, and no validated biomarkers for routine clinical use are established.

The pathophysiology of PCS is incompletely understood. Several non-mutually exclusive mechanisms have been proposed, including viral persistence, with SARS-CoV-2 spike protein and RNA detectable in plasma and diverse tissue reservoirs months after acute infection^4,5^; persistent complement dysregulation and thromboinflammatory signatures^6^; chronic immune dysregulation with T cell activation and autoantibody formation; endothelial dysfunction and microclotting and reactivation of latent herpesviruses^2,6^. The clinical overlap with myalgic encephalomyelitis/chronic fatigue syndrome (ME/CFS), a post-infectious condition triggered by diverse viral pathogens^7^, suggests that PCS may represent a broader phenomenon of infection-triggered chronic disease, reinforcing the need for biomarker-based stratification approaches that can capture shared pathobiological features across post-viral syndromes. A fundamental challenge is the clinical heterogeneity of PCS itself: its symptom spectrum spans neurological, cardiovascular, respiratory, and immunological domains^3^, suggesting that distinct clinical phenotypes may exist and motivating stratified diagnostic approaches^35^.

To date, biomarker discovery for PCS has yielded inconsistent results. A systematic review and meta-analysis of over 20 candidate blood biomarkers, including C-reactive protein, D-dimer, and interleukin-6, found statistically significant but modest elevations with considerable inconsistency across studies^8^, and no single analyte has demonstrated the sensitivity and specificity required for clinical use. Multi-omics approaches have shown greater promise for patient classification^6^, but require costly analytical pipelines impractical for clinical settings.

Extracellular vesicles (EVs) offer a potentially more accessible alternative. EVs are lipid bilayer-enclosed particles released by virtually all cell types, carrying cargo that reflects the physiological and pathological state of their cell of origin^9^. Their surface markers, including tetraspanins, integrins, and lineage-specific molecules, provide information about the identity and activation state of originating cells, enabling minimally invasive liquid biopsy approaches^10,11^. Multiplex bead-based flow cytometry platforms allow simultaneous profiling of dozens of EV-associated surface epitopes^12^. In the context of SARS-CoV-2 infection, EV cargo proteomes have revealed diagnostic potential as severity biomarkers^13^, EVs have been shown to display spike-derived peptides and modulate immune responses^14^, EV surface profiles can differentiate COVID-19 from influenza^15^, and altered EV miRNA signatures have been identified in post-COVID-19 ME/CFS^16^. However, systematic high-throughput EV surface marker profiling specifically in PCS using standardized multiplex platforms has not been performed.

Here, we used bead-based multiplex flow cytometry to profile 37 EV surface markers in plasma and cerebrospinal fluid using two complementary detection strategies. We evaluated whether multivariable EV profiles distinguish COVID^post^ from COVID^reco^ participants under fully nested internal cross-validation and explored their associations with clinical measures and longitudinal change. Participants with non-SARS-CoV-2 post-viral syndromes were included for exploratory comparison. Our results identify an internally cross-validated EV surface-marker pattern associated with COVID^post^ versus COVID^reco^ cohort status.

## Material & Methods

### Recruitment and stratification of participant cohort

Participants were assigned to three cohorts: Post-COVID-19 syndrome (COVID^post^), recovered COVID-19 (COVID^reco^), and participants with post-viral syndrome of non-SARS-CoV-2 aetiology (non-COVID^post^). The participant selection process is summarized in Figure 1. After participant-identity and cohort adjudication, the analysis roster comprised 151 participants: 61 with post-COVID-19 syndrome (COVID^post^), 80 fully recovered participants (COVID^reco^), and 10 participants with non-SARS-CoV-2 post-viral syndromes (non-COVID^post^). Participants were recruited prospectively (COVID^post^ n=47, COVID^reco^ n=80, non-COVID^post^ n=10) at the Department of Neurology, Ulm University Hospital, between March 2023 and November 2024. Of 59 prospectively assessed participants with suspected post-COVID-19 syndrome, 12 were excluded because their symptoms were explained by an alternative diagnosis. To increase sample size, the COVID^post^ cohort was supplemented with 14 retrospective post-COVID-19 syndrome cases from our institutional Biobank at the Department of Neurology, Ulm University Hospital; these retrospective cases provided nine paired CSF/EDTA-plasma samples, two CSF-only samples, and three EDTA-plasma-only samples, so that the COVID^post^ cohort contributed 56 paired CSF/EDTA-plasma samples, two CSF-only samples, and three EDTA-plasma-only samples in total. Among the COVID^reco^ cohort, 50 participants provided paired CSF/EDTA-plasma and 30 provided EDTA-plasma only, and all 10 non-COVID^post^ participants provided paired CSF/EDTA-plasma. Participants with post-COVID-19 syndrome (COVID^post^ n=61) and post-viral syndromes other than PCS (non-COVID^post^ n=10) were recruited from the post-COVID-19 outpatient unit. PCS was diagnosed according to the WHO Delphi consensus criteria^1^ and additionally required a confirmed positive PCR test for SARS-CoV-2 RNA or antigen rapid test against spike S1 antigen during the acute infection phase. All PCS participants presented with persistent symptoms lasting at least 12 weeks after infection that could not be explained by an alternative diagnosis. Diagnosis required comprehensive differential diagnostic work-up according to the presenting symptoms, including cardiological, gastroenterological, rheumatological, psychosomatic, and age-appropriate oncological evaluations (e.g., gynaecological and urological screening). Non-COVID^post^ individuals had a documented viral infection of different origin (e.g. Epstein-Barr virus) confirmed by medical records. For post-viral cases before 2020, SARS-CoV-2 was excluded by temporal criteria; for cases after 2020, a negative antigen rapid test and/or PCR at the time of acute infection was required. COVID-19 recovered controls (COVID^reco^, n=80) were recruited through the neurological emergency department and public advertisements. These individuals had a confirmed prior SARS-CoV-2 infection (positive antigen rapid test or PCR test) but had fully recovered without persistent symptoms. Participants recruited via the emergency department received magnetic resonance imaging (MRI) of the brain and lumbar-(LP) and venipuncture as part of their routine diagnostics. The COVID^reco^ patient-control subgroup included participants evaluated for dizziness, idiopathic intracranial hypertension, peripheral facial nerve palsy, somatoform disorder, tension-type headache, or medically unexplained visual symptoms. Among the participants enrolled through public advertisements, 30 underwent venipuncture only, while the remainder additionally consented to lumbar puncture as part of the study. Exclusion criteria for all groups included severe psychiatric disorders (e.g. schizophrenia or severe episode of depression), and neurological or medical conditions known to affect neurocognitive functioning (e.g. stroke or previously diagnosed neurocognitive disorder). All participants were screened by a trained physician (DKE) for major physical or psychiatric conditions. All prospective COVID^post^ and non-COVID^post^ participants additionally received a detailed neurological and neuropsychological examination, brain MRI, LP, and venipuncture. Neuropsychological testing was performed in 66 participants of the COVID^reco^ cohort who provided consent for this assessment. Twenty-nine COVID^post^ participants were evaluated at baseline and 1-year-follow-up.

**Figure 1:**
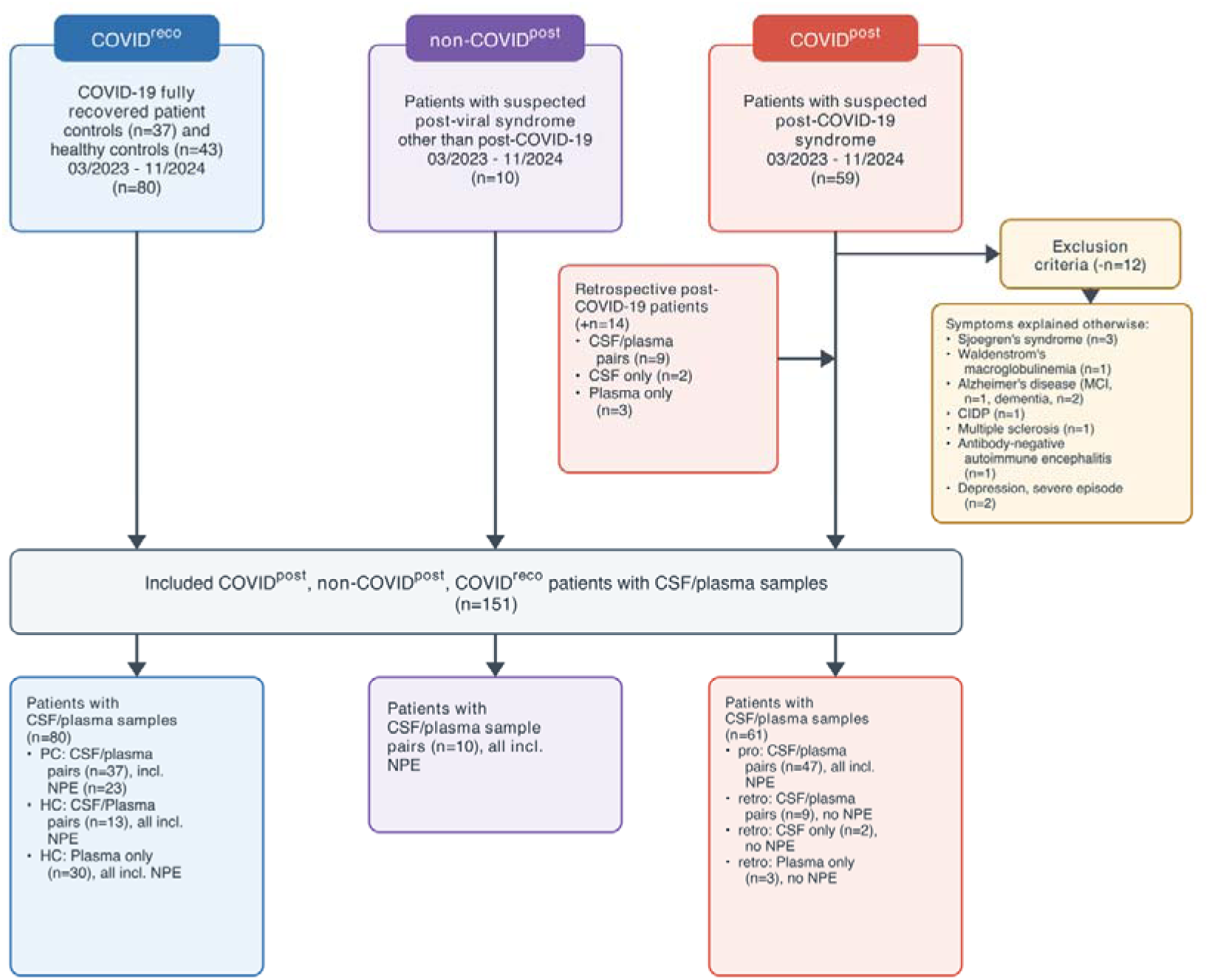
Participant and sample recruitment for the different cohorts. According to the displayed flow chart, participants were divided into three groups: COVID^post^ (post-COVID-19 syndrome (PCS) according to the WHO Delphi consensus) ^1^, non-COVID^post^ (participants with post-viral syndrome other than PCS), and COVID^reco^ (COVID-19 fully recovered controls, n=80). Twelve participants were excluded from the COVID^post^ cohort because their symptoms were explained by an alternative diagnosis, yielding a final analysis roster of 151 participants (COVID^post^ n=61, COVID^reco^ n=80, non-COVID^post^ n=10). The flow chart also depicts the number and type of samples included in the analysis. CSF: cerebrospinal fluid, plasma: EDTA-plasma, PC: patient controls, HC: healthy controls, NPE: neuropsychological examination.

**Figure 2:**
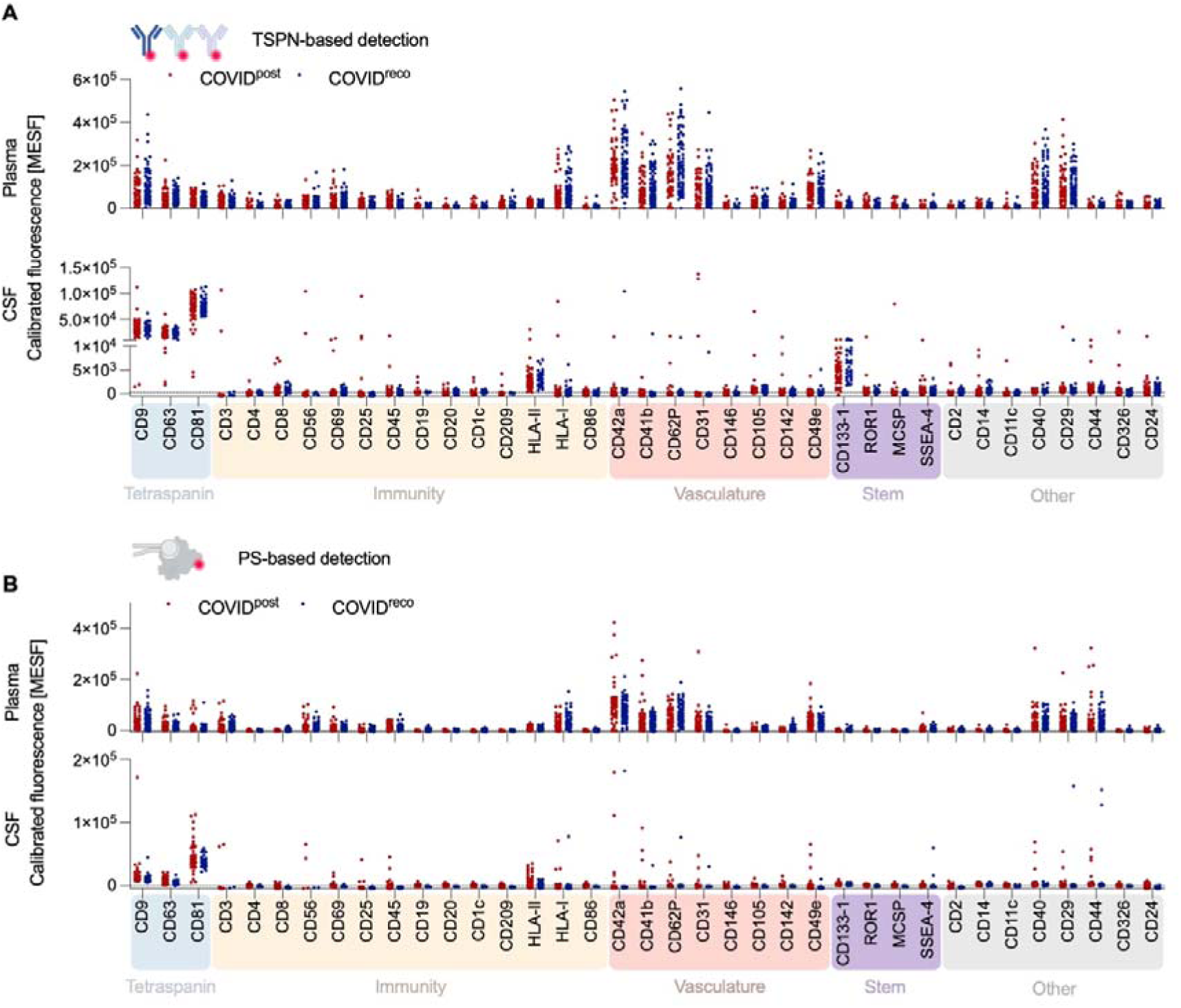
EV surface-marker profiles in post-COVID-19 syndrome (COVID^post^) and COVID-19-recovered controls (COVID^reco^). Extracellular vesicles (EVs) from EDTA plasma and cerebrospinal fluid (CSF) were captured on MACSPlex beads and profiled for 37 surface markers. EV detection was performed using (A) an anti-tetraspanin cocktail targeting CD9, CD63 and CD81 (TSPN) or (B) lactadherin, a phosphatidylserine-binding protein (PS). Fluorescence intensities were converted to molecules of equivalent soluble fluorophore (MESF) using APC calibration for TSPN detection and Alexa Fluor 647 calibration for PS detection. Each dot represents one participant; red: COVID^post^, blue: COVID^reco^. Maximum sample sizes for both detection strategies were plasma n = 52 versus 80 and CSF n = 49 versus 38 (COVID^post^ versus COVID^reco^). All acquired samples were included irrespective of bead count; marker-specific sample sizes were lower where raw fluorescence measurements were missing or could not be calibrated. Markers are grouped by biological function: tetraspanin/EV markers, immune markers comprising T/NK-cell and B-cell/APC subsets, vasculature markers, stem markers, and other markers not assigned to a specific panel. Groups were compared separately for each marker using two-sided Mann–Whitney U tests, with multiple-testing correction performed separately within each 37-marker panel using the two-stage linear step-up procedure of Benjamini, Krieger and Yekutieli (Q = 1%), which did not yield any discoveries; at Q = 5%, PS-detected CD2 was the sole discovery. Dashed lines indicate the LLOQ for APC/TSPN detection (284 MESF) and Alexa Fluor 647/PS detection (820 MESF) as graphical reference lines.

### Neuropsychological assessment

The neuropsychological examination (NPE) comprised testing of different cognitive domains with validated German-language instruments. We evaluated attention (divided attention, alertness, incompatibility) using the German Testbatterie zur Aufmerksamkeitsprüfung (TAP)^17^. We evaluated visual attention and information-processing speed with the Symbol Digit Modalities Test (SDMT)^18^. Verbal fluency was tested with the Regensburger Wortflüssigkeitstest (RWT)^19^, the German version of verbal-fluency measures. Working and verbal short-term memory were assessed using the Digit Span Test from the Wechsler Memory Scale-Revised (WMS-R)^20^. Non-verbal short-term and working memory were also tested with the Block-Tapping Test from the WMS-R^20^. Verbal episodic memory was assessed using the Verbal Learning and Memory Test (VLMT)^21^, and non-verbal episodic memory was tested with the Rey–Osterrieth Complex Figure Test (ROCF)^22^. Cognitive and motor fatigue were assessed using the 20-item Fatigue Scale for Motor and Cognitive Functions (FSMC; five-point Likert scale)^23^. Motor-fatigue scores were categorized as mild (22–26), moderate (27–31), or severe (≥32); cognitive-fatigue scores as mild (22–27), moderate (28–33), or severe (≥34); and total scores as mild (43–52), moderate (53–62), or severe (≥63). For the present dataset, age- and education-corrected normative z-scores were available only for the SDMT^18^. Using the available age- and education-corrected normative scores and the norm-referenced impairment cutoff applied in the clinical export, impaired SDMT performance was observed in 42.6% of the COVID^post^ cohort compared with 7.6% of the COVID^reco^ cohort. The low prevalence of SDMT impairment in the COVID^reco^ group is compatible with the known occurrence and variability of isolated low neuropsychological scores in healthy individuals^25,26^. For the remaining analyses, z-standardization of the 20 cognitive subtests used a 67-control neuropsychological reference set comprising the 66 coded COVID^reco^ participants included in the EV-linked clinical analysis and one additional anonymous healthy control whose neuropsychological data were available for reference calculation but whose EV/roster identifier could not be linked; z-scores were derived from the pooled mean and standard deviation (SD) of this reference set. Consequently, Table 1 and Figure 4 include 66 COVID^reco^ participants. Six composite scores (cs) were calculated by averaging the respective subtest z-scores. For descriptive analyses, performance more than 1 SD below the COVID^reco^ reference mean was operationally labelled as lower relative performance; this study-specific threshold does not by itself establish clinical cognitive impairment^24–26^. The global cognition score was derived from all 20 neuropsychological subtests. The short-term-memory composite (verbal and non-verbal) comprised the forward conditions of the two WMS-R subtests, namely the Digit Span Test and the Block-Tapping Test. The working-memory composite (verbal and non-verbal) was calculated from the corresponding backward versions of these WMS-R subtests. The attention-and-executive-functions composite was obtained by averaging the z-scores of the SDMT, TAP subtests (divided attention, incompatibility, and alertness), and the RWT. The attention-only composite was constructed from the SDMT and TAP subtest z-scores. Finally, the long-term-memory composite (verbal and non-verbal) was based on the VLMT total learning score, VLMT trial 7, VLMT delayed recall, and ROCF delayed recall.

**Table 1:** Demographic and clinical characteristics of the three study cohorts.

| Characteristics | COVID<sup>post</sup><br>(n=47) | COVID<sup>reco</sup><br>(n=66) | non-COVID<sup>post</sup><br>(n=10) | p-value |
| --- | --- | --- | --- | --- |
| Age (y) | 46.98 (33.14-54.11) | 39.69 (31.99-55.00) | 35.49 (26.15-45.27) | 0.27 |
| Sex (f/m), % | (33/14), 70.2%/29.8% | (38/28), 57.6%/42.4% | (6/4), 60.0%/40.0% | 0.40 |
| COVID-19 vaccination at study admission (+/-), n (%) | (44/3), 93.6%/6.4% | (60/6), 90.9%/9.1% | (9/1), 90%/10% | 0.79 |
| Hospitalization during acute COVID-19 (+/-), % | (1/46), 2.1%/97.9% | 0% | 0% | 0.46 |
| Education (y) | 14.00 (13.00-16.00) | 15.00 (13.00-17.00) | 15.25 (12.88-18.25) | 0.40 |
| Time since infection to biosampling (y) | 1.74 (1.05-2.25) | 1.44 (0.67–1.96) | 3.32 (1.21-7.43) | 0.005 |
| FSMC total | 85.00 (81.00-90.00) | 28.00 (23.00-42.25) | 75.00 (69.50-85.25) | < 0.001 |
| - Motor | 43.00 (41.00-46.00) | 14.50 (11.00-20.25) | 37.00 (32.50-45.25) | < 0.001 |
| - Cognition | 43.00 (41.00-46.00) | 14.00 (11.00-22.00) | 39.00 (32.25-43.00) | < 0.001 |
| Global Cognition (cs) | -0.95 (-1.83 to -0.24) | 0.01 (-0.19 to 0.35) | -0.59 (-1.21 to -0.24) | < 0.001 |
| Short-term memory v+nv (cs) | -0.82 (-1.29- -0.02) | 0.01 (-0.56-0.59) | -0.31 (-0.75- -0.05) | < 0.001 |
| Working memory v+nv (cs) | -0.67 (-1.18 to -0.06) | -0.05 (-0.58-0.47) | -0.77 (-1.11- -0.45) | < 0.001 |
| Long-term memory v+nv (cs) | -0.43 (-1.44-0.08) | 0.20 (-0.36-0.57) | -0.51 (-1.55-0.60) | < 0.001 |
| Attention (cs) | -1.22 (-4.83- -0.11) | 0.01 (-0.41-0.49) | -1.03 (-2.23- -0.46) | < 0.001 |
| Attention + executive functions (cs) | -1.02 (-2.67- -0.16) | -0.03 (-0.37-0.32) | -0.70 (-1.46- -0.40) | < 0.001 |

y, years; f/m, female/male; FSMC, Fatigue Scale for Motor and Cognitive Functions; cs, composite score; V+NV, verbal and non-verbal. Values are median (interquartile range) or n (%). P values compare COVID<sup>post</sup> and COVID<sup>reco</sup> using two-sided Mann–Whitney U tests for continuous variables and Fisher’s exact tests for categorical variables.

### CSF collection and laboratory analyses

Cerebrospinal fluid (CSF) and serum were collected either as part of a routine lumbar puncture or voluntarily in 13 COVID^reco^ participants; all procedures were performed by the same physician (DKE). CSF and serum routine diagnostics were conducted according to the guidelines of the German Society for Cerebrospinal Fluid Diagnostics and Clinical Neurochemistry (DGLN) and the German Society of Neurology (DGN)^27^. Briefly, leucocytes and erythrocytes were counted using a Fuchs–Rosenthal chamber. The remaining CSF and serum were centrifuged at 2,000 × g for 10 min, and the supernatant was stored at −30°C for further analyses. CSF lactate was measured photometrically using AU400/AU680 Clinical Chemistry Analyzers (Olympus/Beckman Coulter, Krefeld, Germany). Total protein was measured using a cobas c 503 analyser (Roche Diagnostics; assay reference 08058679 214). Albumin and immunoglobulins G, A, and M were measured in CSF and serum using a nephelometer (Atellica NEPH 630 System; Siemens Healthcare GmbH, Erlangen, Germany). Oligoclonal IgG bands in paired CSF and serum were assessed by isoelectric focusing as part of routine clinical diagnostics, consistent with recommended CSF analysis standards^28^.

Q_Alb_ represents the integrity of the blood-CSF barrier and was calculated as CSF/serum ratio of albumin. The age-dependent upper limit was calculated using Reiber’s formula^40^. Additionally, quantitative intrathecal immunoglobulin G-/A-/M-synthesis was also assessed using CSF/serum ratios (Q_Ig_total_) and taking the individual Q_Alb_ into account with respect to their upper limits (Q_lim_). Qlim for each immunoglobulin was calculated using Reiber’s formulas, and intrathecal synthesis of the respective immunoglobulin was assumed when QIg_total exceeded the corresponding Qlim^30^. Pathogen-specific antibody-indices (AIs) were calculated using pathogen-specific ratios for CSF/serum IgG (Q_spec_). AIs were built by dividing Q_spec_ by Q_Ig_total_ for Q_Ig_total_ < Q_lim_ and Q_spec_/Q_lim_ for Q_Ig_total_ exceeding Q_lim_^29^. Pathogen-specific IgG synthesis was assumed for AI ≥ 1.5.

### Blood collection and platelet-free plasma preparation

Venous blood was collected by venipuncture concurrently with LP, if performed, using a 20G Safety-Multifly needle (Ref# 85.1637.235, Sarstedt) with brief tourniquet application. A volume of 7.5 mL was drawn into K3-EDTA S-Monovettes (Ref# 01.1605.001, Sarstedt) and gently inverted three times immediately after collection. Within 30 minutes of phlebotomy, platelet-free plasma (PFP) was prepared following the double-centrifugation protocol recommended by the International Society on Thrombosis and Haemostasis (ISTH). Briefly, samples were centrifuged at 2,500 × g for 15 minutes at room temperature without brake. The supernatant plasma was carefully collected leaving approximately 10 mm above the cell pellet and transferred to a new 15 mL conical tube. The centrifugation step was repeated under identical conditions (2,500 × g, 15 min, RT, no brake), and the resulting double-centrifuged plasma was again harvested 10 mm above the pellet. PFP was aliquoted (5 × 250 µL per participant) and stored at −80°C until analysis. All plasma samples underwent a single freeze–thaw cycle before analysis.

### Bead-assisted flow cytometric profiling of EV surface markers

EV-containing samples were subjected to bead-based multiplex EV analysis (MACSPlex Exosome Kit, human, Miltenyi Biotec) as previously described with adaptations^31^. Briefly, samples were diluted 1:1 in MACSPlex buffer (per assay, 5 μl capture beads and 30 μl plasma or CSF mixed with 30 μl buffer) and incubated overnight with MACSPlex Exosome Capture Beads on an orbital shaker at 450 rpm at room temperature. Beads were washed with MACSPlex buffer, and most of the supernatant was aspirated. For staining of captured EVs, a cocktail of APC-conjugated anti-CD9, anti-CD63, and anti-CD81 detection antibodies (Tetraspanins (TSPN); Miltenyi Biotec; #130-108-813; 5 µl each) or alternatively for detection of PS, 0.5 μg LA-A647 (Haematologic Technologies, #BLAC-ALEXA647) were added to the tube followed by incubation at 450 rpm for 1 h at room temperature in the dark. Next, the samples were washed twice with MACSPlex buffer and liquid removed before resuspension in 150 µl MACSPlex buffer. Samples were then transferred to a V-bottom 96-well microtiter plate (Thermo Scientific) and analysed by flow cytometry using a Cytoflex LX (Beckman Coulter). CytExpert 2.3 (Beckman Coulter) was used to analyse flow cytometric data (see Supplementary Information for gating strategy). Median fluorescence intensities were extracted for the 37 analyte capture-bead populations; the two MACSPlex kit control populations were not included as model features. Fluorescence was calibrated to molecules of equivalent soluble fluorophore (MESF) using calibration beads (Bangs Laboratories Quantum). Calibration curves were acquisition- and fluorophore-specific and were fitted in log10 fluorescence–log10 MESF space using the non-zero fluorescence standards. Sample and matched buffer-only blank fluorescence values were converted to MESF separately using the corresponding acquisition- and fluorophore-specific curve. For each acquisition, stain, and marker, valid calibrated blank-replicate MESF values were averaged and then subtracted from the calibrated sample MESF. Non-positive compensated blank fluorescence values, which cannot be log-transformed, were assigned the physical lower bound of 0 MESF before blank averaging. Signed blank-subtracted values, including values below zero, were retained without replacement by any lower limit of quantification (LLOQ). Technical replicates acquired within the same run were averaged. The graphical LLOQs, used only as reference lines, were 284 MESF for APC/TSPN detection and 820 MESF for Alexa Fluor 647/PS detection. Consistent with MISEV2023 technique-specific terminology, this workflow was used as a direct bead-based affinity-capture profiling assay rather than as a procedure for EV purification or single-particle characterization^9^. The fluorescence readout therefore represents bead-associated material carrying both the capture epitope and the TSPN or PS detection target; conventional purity metrics and immunoblotting of an isolated EV fraction were not generated by this workflow, and contributions from non-vesicular co-associated material cannot be excluded^9^.

### Nanoparticle tracking analysis

Particle concentration and size distributions of nanoparticles in plasma and CSF were measured by nanoparticle tracking analysis on a ZetaView TWIN (Particle Metrix), as previously described^31^. For concentration and size measurements, samples were diluted in particle-free PBS, and the scattering particles were recorded under the following settings: fixed temperature of 25 °C, 11 positions, 1 cycle, sensitivity 85–90, shutter 100, 15 fps, 2 s per position, and 3–5 measurements per sample. The resulting videos were analysed with ZetaView Analyze 08.05.05 SP2 and 08.05.12 SP1. The measurement chamber was thoroughly flushed with particle-free PBS between samples.

### Machine learning-based classification of EV surface marker profiles

To distinguish COVID^post^ from COVID^reco^, plasma profiles were analysed separately for TSPN and PS detection. Each matrix comprised the same 132 participants (COVID^post^ n=52, COVID^reco^ n=80) and 37 signed, blank-subtracted MESF features; COVID^post^ was the positive class. We used L1-regularized logistic regression with stratified nested cross-validation. The outer evaluation comprised five-fold cross-validation repeated five times using seeds 42–46. Within every outer-training set, features with more than 50% missingness were removed, remaining missing values were imputed using training-set medians, and features were standardized using training-set means and standard deviations. These fitted transformations were then applied to the corresponding held-out data. No imputation, scaling, regularization tuning, or feature selection used observations from the outer test fold. Regularization strength was selected by five-fold stratified inner cross-validation from 10 logarithmically spaced C values between 10 and 10 , maximizing mean inner-fold ROC AUC, with the smaller C selected in the event of a tie. Models used the SAGA solver with an L1 penalty, max_iter=100,000, and tolerance 10 . The All model used all 37 candidate features, with sparsity determined by the fitted L1 coefficients. Two adaptive reduced models were additionally evaluated. Within each outer fold, a full LASSO selector was refitted within the inner-training partitions, and markers with nonzero coefficients were ranked by absolute coefficient magnitude. Candidate prefix sizes and the reduced-model regularization strength C were evaluated using inner-validation data. The CV-selected model used the panel size with the highest mean inner-cross-validation AUC, whereas the CV-1SE model used the smallest panel size whose mean inner-cross-validation AUC was within one standard error of the optimum. Both marker identity and panel size were therefore selected using training data only and could differ between outer folds; these are adaptive selection procedures rather than fixed biomarker panels. The four prespecified biological marker sets were immunity (CD3, CD4, CD8, CD19, CD20, CD1c, CD209, HLA-DR/DP/DQ, HLA-ABC, CD86, CD56, CD69, CD25, CD45), vasculature (CD42a, CD41b, CD62P, CD31, CD146, CD105, CD142, CD49e), stem (CD133-1, ROR1, MCSP, SSEA-4), and other (CD2, CD14, CD11c, CD40, CD29, CD44, CD326, CD24). These sets were analysed using the same fold-local preprocessing and regularization tuning. For each repeat, every participant received one out-of-fold predicted probability from a model that had not been trained on that participant. Figure 3C reports the arithmetic mean of the five repeat-specific out-of-fold AUCs; plotted ROC curves are pointwise means of the five repeat-specific out-of-fold ROC curves. Figure 3D scores are the participant-wise means of the five out-of-fold predicted probabilities. Uncertainty in mean repeat AUCs was quantified using 10,000 cohort-stratified participant-level bootstrap resamples (seed 20260826). COVID^post^ and COVID^reco^ participants were sampled with replacement within cohort, retaining each participant’s vector of five repeat-specific out-of-fold predictions as a unit. For each bootstrap resample, AUC was calculated separately in every repeat and then averaged; percentile 95% CIs were defined by the 2.5th and 97.5th percentiles. These CIs are conditional on the fitted cross-validation predictions and capture participant-sampling uncertainty but not full model-refitting or external-validation uncertainty. All analyses were performed in Python 3.11.2 (NumPy 2.2.6, pandas 2.3.3, SciPy 1.15.3, scikit-learn 1.7.2, and statsmodels 0.14.5).

**Figure 3:**
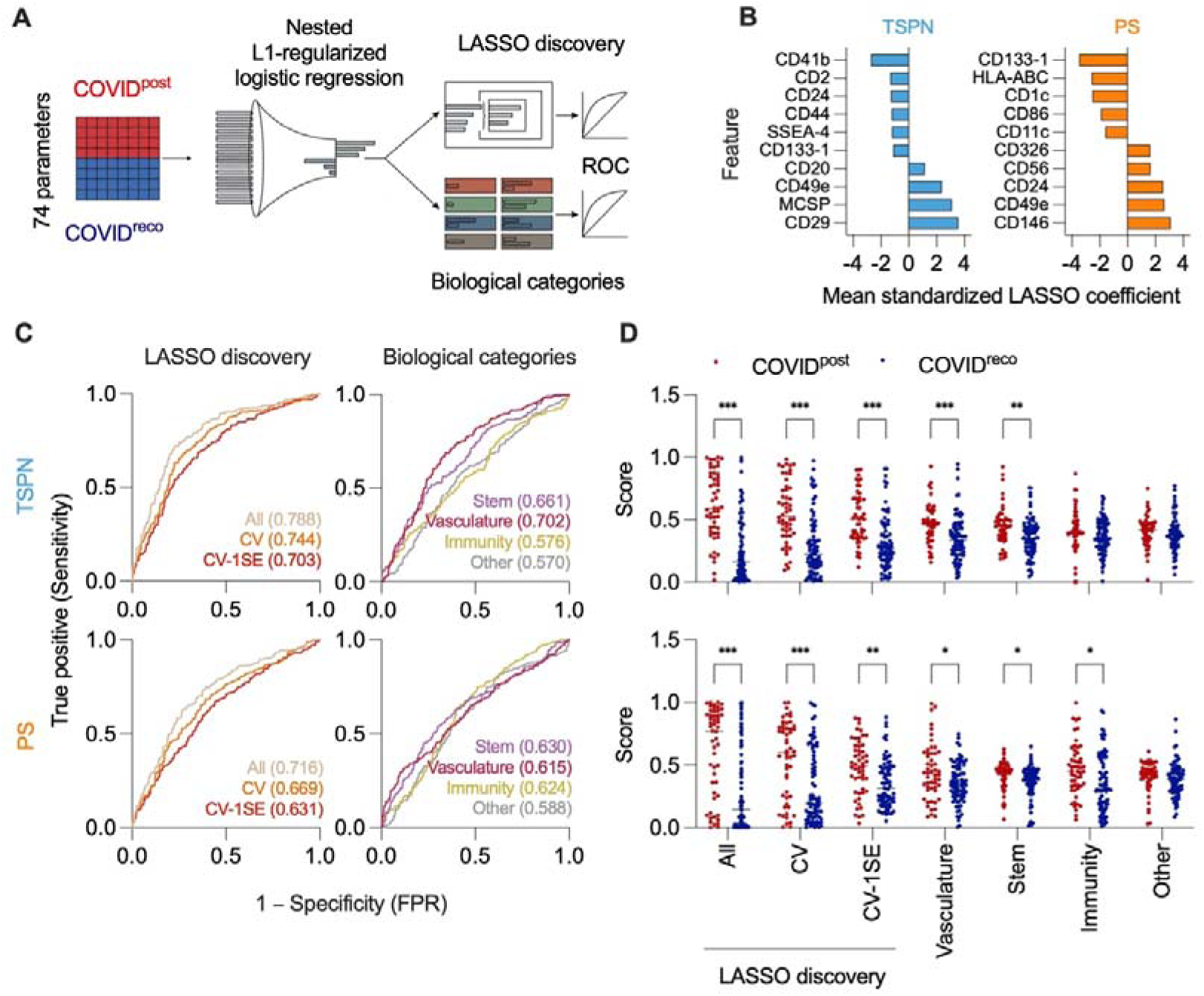
LASSO-based classification of post-COVID-19 syndrome using plasma EV surface-marker profiles. (A) Schematic of the strictly nested analysis workflow. TSPN- and PS-detected profiles were analysed separately using L1-regularized logistic regression. Data-driven analyses comprised a full 37-marker input model (All), a reduced model whose panel size maximized inner-cross-validated AUC (CV-selected), and a parsimonious model using the smallest panel size within one standard error of the optimum (CV-1SE). Preprocessing, feature ranking, regularization tuning, and panel-size selection were performed using training data only. Fixed biology-based panels comprised Vasculature, Stem, Immunity, and Other markers. (B) The ten markers with the largest absolute mean standardized LASSO coefficients across the 25 outer-training fits for TSPN and PS. Positive coefficients indicate a higher predicted probability of COVID^post^; this cross-fold summary is descriptive and does not define a fixed reduced panel. (C) Receiver operating characteristic curves from five repeats of stratified five-fold outer cross-validation. Values in parentheses are mean repeat AUCs calculated from out-of-fold predictions. Corresponding participant-bootstrap 95% CIs are reported in Table S2. Because selection was repeated within the training data, the size and composition of the CV-selected and CV-1SE panels could vary across folds. (D) Participant-level classification scores obtained by averaging each participant’s five out-of-fold predicted probabilities. Red: COVID^post^ (n = 52); blue: COVID^reco^ (n = 80). Group comparisons were performed using two-sided Mann–Whitney U tests with Holm– Šidák correction across the seven displayed panels separately within each detection strategy. *adjusted p < 0.05, **adjusted p < 0.01, ***adjusted p < 0.001.

**Figure 4:**
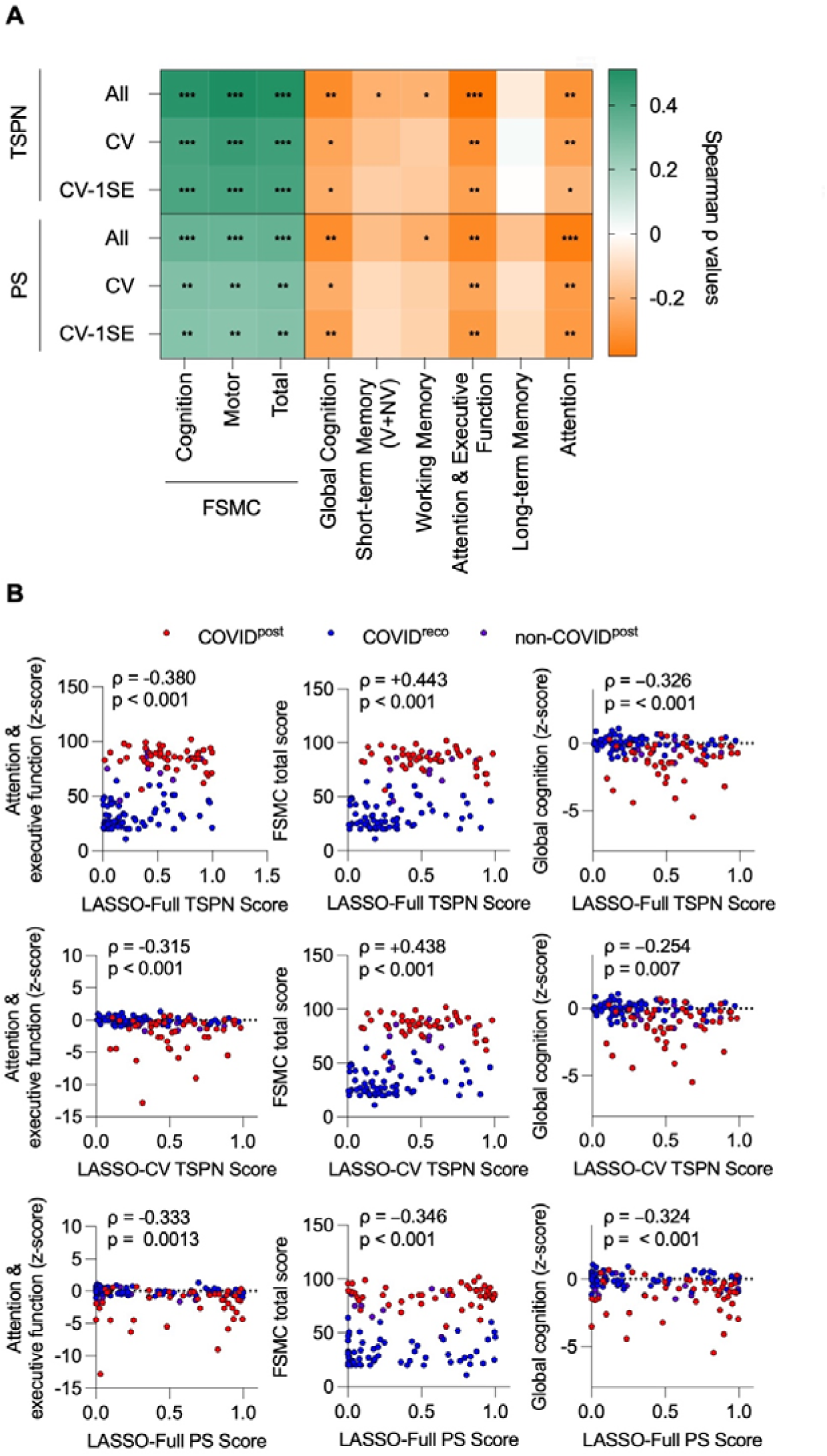
**Association of EV-based classification scores with clinical parameters. (**A) Spearman correlations between nested cross-validated plasma EV-panel classification scores and clinical parameters for TSPN (top) and PS (bottom) detection in pooled COVID^post^ and COVID^reco^ participants (n = 113; COVID^post^ n = 47, COVID^reco^ n = 66). Heatmap rows are TSPN All, TSPN CV-selected, TSPN CV-1SE, PS All, PS CV-selected, and PS CV-1SE; the CV-selected and CV-1SE panels were reselected within the training folds of the nested cross-validation. Color indicates Spearman _ρ_. P values were corrected across the 54 displayed correlations using the Benjamini–Hochberg procedure; *q < 0.05, **q < 0.01, ***q < 0.001. V+NV, verbal and non-verbal; FSMC, Fatigue Scale for Motor and Cognitive Functions. (B) Representative scatter plots for TSPN All (top row), TSPN CV-selected (middle row), and PS All (bottom row) against attention and executive function (left), FSMC Total (centre), and global cognition (right). Higher EV classification scores indicate a greater predicted probability of COVID^post^. Each dot represents one participant; red, COVID^post^ (n = 47); blue, COVID^reco^ (n = 66); purple, other post-viral syndrome (n = 9). Annotations show Spearman _ρ_ and the Benjamini–Hochberg-adjusted q value from the complete 54-test family, calculated using COVID^post^ and COVID^reco^ participants only. Purple points were excluded from all correlation statistics because of the small sample size and are shown for visual reference only.

### Ethics approval and participant consent

The study was conducted in accordance with the Declaration of Helsinki and approved by the Ethics Committee of Ulm University (approval number 16/23 from 16^th^ March 2023). All participants provided written informed consent prior to enrolment. They also consented to the post-processing of the data, e.g. for publications.

### Statistics

Statistical analyses were performed using GraphPad Prism V.10.2.2 (GraphPad Software, La Jolla, CA) or Python 3.11.2 using NumPy 2.2.6, pandas 2.3.3, SciPy 1.15.3, and scikit-learn 1.7.2 for macOS. For details on machine learning, see separate section. As EV surface marker intensities are not expected to follow normal distributions, all group comparisons were performed using non-parametric tests (Mann-Whitney U for two groups, Kruskal-Wallis for multiple groups). We separated the three cohorts COVID^post^, non-COVID^post^, and COVID^reco^ according to the existence of persistent symptoms after a COVID-19 infection and according to the WHO Delphi consensus1. Testing for normal distribution was performed using the Shapiro-Wilk test. All tests were two-sided. Categorical variables are reported as counts and percentages and continuous variables as medians and interquartile ranges. Group comparisons used Mann–Whitney U, Kruskal–Wallis, or Fisher’s exact tests as appropriate. Missing observations were omitted on a test-wise basis for univariate and correlation analyses; missing EV features in machine learning were handled only by the fold-local imputation procedure described above. Multiplicity families were defined by figure. For Figure 2, the two-stage linear step-up procedure of Benjamini, Krieger and Yekutieli was applied separately within each 37-marker assay and compartment family at the stringent Q = 1% threshold; for transparency, discoveries at the conventional Q = 5% threshold were also reported. For Figure 3D, two-sided Mann–Whitney U tests were corrected using the Holm– Šidák procedure across the seven displayed models, separately within TSPN and PS detection, and are reported as adjusted p values. Benjamini–Hochberg correction was applied across the 54 Figure 4A correlations and across the six Figure 5 paired comparisons. Statistical significance was defined using the figure-specific adjusted thresholds described above; unadjusted tests used p < 0.05. The study follows the STROBE guidelines for cross-sectional (prospective) studies. Clinical associations were assessed using each participant’s mean of five nested-cross-validated out-of-fold COVID^post^ probabilities, joined to clinical data by full canonical participant identifier. Primary heatmap analyses comprised six EV-score procedures × nine clinical measures in 113 participants with clinical matching (COVID^post^ n=47, COVID^reco^ n=66) and used two-sided Spearman correlations on pairwise-complete observations pooled across the two cohorts. Benjamini–Hochberg correction was applied across the 54 displayed tests. Within-COVID^post^ and within-COVID^reco^ correlations, together with rank-based partial Spearman correlations controlling for cohort, were computed as sensitivity analyses to distinguish pooled diagnostic-group concordance from within-cohort severity associations. Scatter plots display Spearman ρ calculated in the pooled COVID^post^/COVID^reco^ cohort together with the same Benjamini–Hochberg-adjusted q values from the complete 54-test family, rather than separate unadjusted p values. Nine participants with other post-viral syndromes were displayed descriptively but excluded from all correlation statistics. The longitudinal analysis comprised 29 COVID^post^ participants with a verified biological follow-up and a baseline included in the final Figure 3 model population. For each cross-validation repeat, the follow-up sample was scored using the exact outer-fold model whose held-out test fold contained that participant’s baseline. Thus, the same model, trained without either measurement from that participant, generated the baseline out-of-fold and follow-up probabilities, including identical fold-specific preprocessing, feature selection, and regularization. Baseline and follow-up probabilities were averaged across the five repeats. Paired differences were assessed using two-sided Wilcoxon matched-pairs signed-rank tests, with Benjamini–Hochberg correction across the six displayed panels.

**Figure 5:**
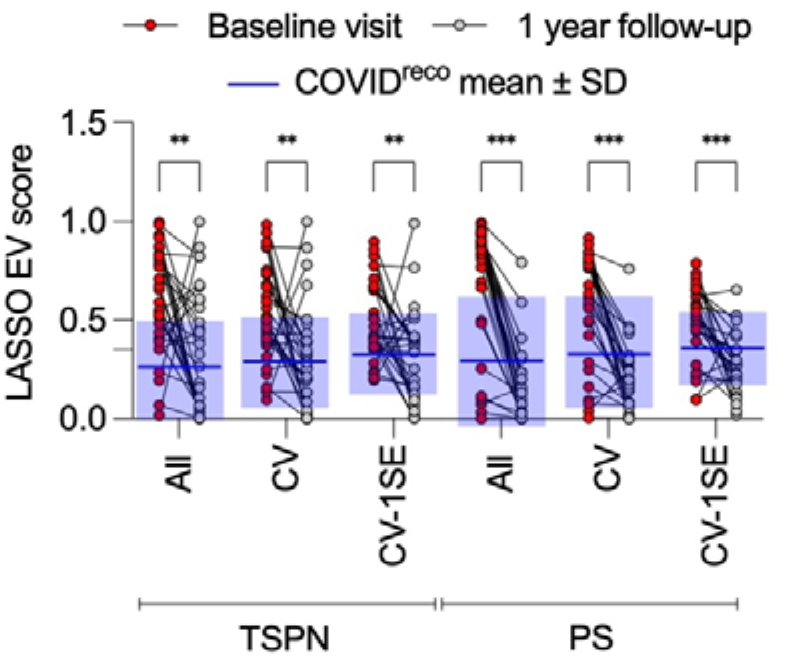
EV-based classification scores decrease at one-year follow-up. Paired cross-fitted EV classification scores at baseline (red) and one-year follow-up (grey) are shown for 29 participants with post-COVID-19 syndrome (48% of the COVID^post^ cohort) across six data-driven models: TSPN All, TSPN CV-selected, TSPN CV-1SE, PS All, PS CV-selected, and PS CV-1SE. ‘All’ denotes the L1-regularized model using all 37 markers; ‘CV-selected’ denotes the reduced model size that maximized mean inner-cross-validation AUC within each outer-training fold; ‘CV-1SE’ denotes the smallest model within one standard error of the best inner-cross-validation AUC. Each connected pair represents the same participant at both time points. Within each cross-validation repeat, the follow-up sample was scored only with the outer model whose test fold contained that participant’s baseline sample; displayed values are means across five repeats. Blue horizontal lines indicate the mean out-of-fold score of COVID^reco^ controls for the corresponding model (n = 80). Baseline and follow-up scores were compared using two-sided Wilcoxon matched-pairs signed-rank tests with Benjamini–Hochberg correction across the six displayed comparisons; *q < 0.05, **q < 0.01, ***q < 0.001.

## Results

### Demographical and clinical data of the cohorts

We studied 61 COVID^post^ participants, 80 COVID^reco^ participants, and 10 non-COVID^post^ participants (Figure 1). Participants provided EDTA plasma and/or CSF as summarized in Figure 1, together with neuropsychological evaluation (NPE) and patient-reported fatigue assessment using the FSMC where available. The prospectively characterized clinical subsets are summarized in Table 1. Overall, COVID^post^ participants showed greater fatigue and poorer performance across several cognitive domains than COVID^reco^ participants, while the small non-COVID^post^ cohort showed a broadly similar clinical phenotype. Exact cohort summaries and statistical comparisons are reported in Table 1. A comprehensive listing of all symptoms observed in the COVID^post^ and non-COVID^post^ participant groups is provided in Table S1 of the Supplementary Material.

In order to evaluate changes in the surface-exposed proteins in circulating EVs of COVID^post^ participants, we performed surface epitope profiling of circulating extracellular vesicles from plasma and CSF of COVID^post^ participants and COVID^reco^ controls using a bead-based flow cytometry setup with an immuno-oncology-oriented marker panel. Here, we analysed EVs captured by a total of 37 antigens, which were then detected by the conventional anti-Tetraspanin (TSPN) cocktail (targeting CD9, CD63 and CD81, Figure 2A) and additionally, using a phosphatidylserine-binding protein to detect a broader EV subset independently of tetraspanins (Figure 2B). For all assays, we used a fixed volume of plasma and CSF as input, as no significant differences were measured in plasma particle levels in NTA (Figure S1) and CSF particle levels were generally low (most samples < 10^10^ particles/ml). Notably, NTA revealed a higher particle concentration in COVID^post^ than COVID^reco^ CSF (p<0.001) and a modestly larger median particle diameter in COVID^post^ plasma (p=0.012); plasma particle concentration and CSF median diameter did not differ significantly.

Most EV surface-marker signals showed broad and overlapping distributions between COVID^post^ and COVID^reco^. Using a stringent 1% false-discovery-rate threshold, no individual marker met the criterion for statistical discovery in either assay (Figure 2). At the conventional 5% threshold, PS/LA-detected CD2 was the sole discovery in plasma (P < 0.001, q = 0.03); no other marker met this threshold. Additional PS/LA markers, including CD105, CD133-1, CD142, SSEA-4, CD19, CD1c, CD69, CD62P and CD11c, showed nominal differences in the same direction (P = 0.006–0.024; q approximately 0.09). In the TSPN assay, the strongest nominal differences were observed for CD62P (P = 0.003, q = 0.10), CD2 (P = 0.006, q = 0.11), CD4 and CD11c (both P approximately 0.01, q = 0.13). Further, there were no significant differences between COVID^post^ and COVID^reco^ in any marker with both stains for CSF samples (Figure 2A,B). It must be noted that this is also influenced by the relatively low concentration of EVs in CSF compared to plasma, meaning that only marker subpopulations with higher abundance can be reliably detected. In CSF, many marker signals were close to or below assay background, limiting the statistical power to detect group differences.

To further explore CSF-derived EVs, we employed a neurology-focused capture bead panel targeting 37 markers enriched for nervous system cell-derived EVs (Supplementary Figure S2). While this panel yielded more quantifiable markers (24/37 above the LLOQ in >50% of samples), including neurology-specific markers such as CX3CR1, Podoplanin, O4, CD11b, CD107a, and CD68, none showed significant differences between COVID^post^ and COVID^reco^ groups, and 7/37 markers remained at or below background. These findings suggest that the overall EV concentration in unconcentrated CSF is insufficient for reliable bead-based surface profiling, regardless of the capture panel used. Together with the considerable interindividual spread observed for most markers, these results indicate limited discriminatory power for individual EV markers.

This motivated a multivariable analysis to test whether combinations of markers provide greater discriminatory information than any single marker (Figure 3). For this, we trained an L1-regularized (LASSO) logistic regression classifier on the full marker profile to identify the most discriminative markers between COVID^post^ and COVID^reco^ participant EVs and to evaluate whether a reduced subset of markers retained discriminative ability in a data-driven manner (Figure 3A). In parallel, we applied LASSO-optimized weighting within biology-based marker panels reflecting functional categories (as in Figure 2: vasculature, stem, immunity, and other). In addition to the LASSO-based feature discovery across all markers, this allowed us to assess whether specific biological EV subpopulations carry a PCS-associated signature (Figure 3A, Table S2, Supplementary Material). Classification performance was evaluated using stratified nested five-fold cross-validation repeated five times, with imputation, standardization, regularization tuning, and feature selection performed on the training data within each cross-validation fold. Plasma TSPN and PS datasets were analysed separately, each comprising 37 EV surface markers.

Figure 3B summarizes mean standardized coefficients across the 25 outer-training models rather than a single fixed marker panel. For TSPN detection, the five largest mean absolute coefficients were observed for CD29, MCSP, CD41b, CD49e, and CD2. Their mean directions were positive for CD29, MCSP, and CD49e and negative for CD41b and CD2. For PS detection, the corresponding markers were CD133-1, CD146, CD49e, HLA-ABC, and CD24, with negative coefficients for CD133-1 and HLA-ABC and positive coefficients for CD146, CD49e, and CD24. Markers from several functional categories contributed to both models. The CV-selected and CV-1SE procedures reselected markers within the training folds, so this cross-fold summary does not define a fixed reduced panel.

Classification performance was assessed as the mean of the five repeat-specific out-of-fold AUCs (Figure 3C). The full 37-marker TSPN model achieved a mean nested cross-validated AUC of 0.788 (95% CI 0.715–0.852), indicating moderate discrimination. Two fold-adaptive reduced models were also evaluated. Within each outer-training fold, the CV-selected procedure chose the marker subset that maximized AUC in the inner cross-validation, whereas the CV-1SE procedure selected the smallest subset whose inner-cross-validated AUC was within one standard error of the maximum. Marker identities and panel sizes were therefore reselected independently within every outer-training fold and do not represent fixed marker signatures. The adaptive TSPN CV-selected and CV-1SE models achieved AUCs of 0.744 and 0.703, respectively. The full PS model achieved a mean AUC of 0.716 (95% CI 0.636–0.792), with lower performance for its CV-selected and CV-1SE models (AUCs 0.669 and 0.631). The full TSPN model had a numerically higher AUC than the full PS model; however, the paired bootstrap difference was 0.072 (95% CI −0.024 to 0.166), so this analysis did not establish superiority between the detection strategies. The reduced performance and variable composition of the adaptive models are consistent with discriminative information being distributed across multiple markers rather than captured by a stable compact panel. Across outer folds, the CV-selected panel size had a median of 10 markers (range 3–26) for TSPN and 23 markers (range 1–32) for PS, whereas the CV-1SE panel size had a median of 7 markers (range 2–20) for TSPN and 11 markers (range 1–22) for PS.

The full models therefore performed best, and no stable compact panel emerged. Among the prespecified biological categories, the vasculature-associated TSPN panel showed the greatest discrimination (AUC 0.702), approaching that of the full TSPN model, followed by the stem panel (AUC 0.661), while the immunity and other TSPN panels performed close to chance (AUCs 0.576 and 0.570). The PS biological-category panels showed modest discrimination (vasculature 0.615, stem 0.630, immunity 0.624, other 0.588). The performance of the vasculature-associated TSPN panel supports a possible contribution from this marker group, although the analysis does not establish a causal vascular mechanism.

The principal classification pattern was preserved in post-hoc robustness analyses. Restricting COVID^post^ to the 47 prospectively recruited participants yielded mean repeat AUCs of 0.780 for TSPN and 0.696 for PS detection. In the clinically characterized subset, the full-panel score remained associated with COVID^post^ status after adjustment for time from infection to biosampling, age, sex, and vaccination status at study admission (TSPN: odds ratio per SD 4.16, 95% CI 2.36–7.32, p<0.001; PS: odds ratio per SD 2.25, 95% CI 1.46–3.48, p<0.001).

Participant-level classification scores (mean out-of-fold predicted probability of post-COVID status) separated COVID^post^ from COVID^reco^ participants for most panels (Figure 3D). For TSPN detection, the All, CV-selected and CV-1SE models, together with the vasculature panel, showed the clearest separation (adjusted p < 0.001), and the stem panel remained significant (adjusted p < 0.01), whereas the immunity and other panels did not reach significance. For PS detection, the All and CV-selected models separated the groups most clearly (adjusted p < 0.001), the CV-1SE model reached adjusted p < 0.01, and the vasculature, stem and immunity panels reached adjusted p < 0.05, while the other panel did not. These score comparisons are descriptive secondary analyses of the cross-fitted probabilities; the nested cross-validated ROC AUCs (Figure 3C) remain the primary evidence of classification performance.

We next examined associations between EV classification scores and fatigue and neuropsychological measures. Strikingly, across the pooled COVID^post^ and COVID^reco^ population, EV classification scores were associated with the clinical phenotype. The full TSPN score correlated positively with FSMC Total (ρ = 0.501, q = 4.17 × 10 ) and negatively with attention and executive function (ρ = −0.380, q = 1.76 × 10 ) and global cognition (ρ = −0.326, q = 0.00141). The TSPN CV-selected score retained the same pattern with somewhat smaller effect sizes (FSMC Total: ρ = 0.437, q = 1.37 × 10 ; attention and executive function: ρ = −0.315, q = 0.00204; global cognition: ρ = −0.255, q = 0.0107). The full PS score showed moderate associations in the same directions (FSMC Total: ρ = 0.346, q = 0.000778; attention and executive function: ρ = −0.333, q = 0.00113; global cognition: ρ = −0.324, q = 0.00150). The directions of all nine representative associations were consistent across all five cross-validation repeats. No association remained significant after correction when examined within COVID^post^ or COVID^reco^ separately, and none remained significant in partial Spearman analyses controlling for cohort, indicating that while these scores separate the cohorts, they do not correlate with severity of clinical phenotypes within the cohorts.

Among 29 COVID^post^ participants with paired baseline and approximately one-year follow-up plasma samples, classification scores decreased in all six models after correction for multiple comparisons. Median changes in follow-up minus baseline score were −0.215 for TSPN All (q = 0.0038), −0.246 for TSPN CV-selected (q = 0.0031), −0.210 for TSPN CV-1SE (q = 0.0031), −0.654 for PS All (q < 0.001), −0.359 for PS CV-selected (q < 0.001), and −0.229 for PS CV-1SE (q < 0.001). Scores decreased in 23 of 29 participants in each TSPN model, in 27 of 29 participants in the PS All and PS CV-selected models, and in 24 of 29 participants in the PS CV-1SE model. These findings show that both TSPN- and PS-based EV profiles shifted toward lower COVID^post^ classification scores at follow-up, consistent with a potentially transient component of the EV-associated changes; however, their relationship to clinical trajectories requires further longitudinal study.

## Discussion

Using two EV detection strategies, we found that no individual plasma marker met a 1% false-discovery-rate threshold, whereas a distributed multivariable EV surface pattern moderately distinguished COVID^post^ from COVID^reco^ participants under fully nested internal cross-validation. The full TSPN and PS models achieved mean out-of-fold AUCs of 0.788 and 0.716, respectively, and the discriminatory signal was distributed across multiple markers rather than concentrated in any single epitope. This represents internally cross-validated exploratory discrimination, however not yet an externally validated biomarker. Classification scores were associated with fatigue and cognition across the pooled diagnostic groups and decreased in a subset with paired one-year samples. These findings support further investigation of EV surface profiling but do not establish a validated diagnostic biomarker, within-PCS severity measure, or marker of disease resolution.

At the individual-marker level, TSPN-detected CD62P and PS-detected CD2 produced the strongest nominal signals and both were lower in COVID^post^. PS-detected CD2 met the conventional 5% false-discovery-rate threshold but not the stringent 1% threshold; CD62P met neither adjusted threshold. CD62P is platelet-associated^34^, whereas CD2 is associated with lymphoid cells; however, bead-capture epitope signals do not necessarily establish the cellular origin or mechanism of the observed differences. The assay also cannot distinguish a change in the number of marker-positive EVs from a change in surface-epitope density. The broad overlap between groups indicates that neither marker is suitable as a standalone classifier.

The full TSPN model had a numerically higher mean AUC than the full PS model (0.788 versus 0.716), but the paired bootstrap CI for the difference included zero and therefore did not establish superiority of one detection strategy. Tetraspanin- and PS-based reagents compare overlapping but non-identical EV populations^31^, and both carried some discriminatory information. Using the same panel of capture beads, evaluating different detection strategies based on lipids or other PCS-associated candidate biomarkers may provide further improvements, which is subject to future work.

In CSF, the majority of markers fell below the limit of quantification with both detection strategies, and a dedicated neurology-focused capture panel did not overcome this (Figure 2, Figure S2). CSF is an attractive compartment because it should be comparatively enriched for brain-derived EVs that are directly relevant to the cognitive phenotype of PCS; however, putative brain-derived EV populations remain difficult to isolate and detect even with dedicated approaches^36^. Bead-based multiplex cytometry of unconcentrated CSF simply lacks the required sensitivity. Pre-concentration of CSF EVs or ultrasensitive single-vesicle detection methods will be needed before the CSF compartment can be exploited for surface-marker profiling. Considering the limited volume of CSF collected during routine lumbar puncture, higher-sensitivity single-vesicle methods, such as nano-flow cytometry, may be required to develop CSF EV-based biomarkers.

The performance of the full TSPN model suggests that group-associated information is distributed across the EV surface profile, which is in line with expectations based on the heterogeneity of PCS. Its nested cross-validated AUC of 0.788 is encouraging for an exploratory and comparatively accessible assay, but it is not directly comparable with externally validated multi-omic classifiers developed in different cohorts. The adaptive CV-selected and CV-1SE models achieved lower AUCs (0.744 and 0.703 for TSPN; 0.669 and 0.631 for PS), and both their sizes and their marker identities varied across outer folds. The full models performed best, and no fixed compact diagnostic panel was validated.

The clinical correlations require careful interpretation. Although pooled COVID^post^-COVID^reco^ correlations survived correction, none did so within either cohort alone, and none remained significant in rank-based partial correlations controlling for cohort. The EV scores therefore covary with clinical differences separating the diagnostic cohorts but cannot currently be interpreted as continuous severity markers within PCS. Nine analysable participants with other post-viral syndromes were displayed for exploratory visual comparison but were not included in the formal correlation analyses. Their distributions overlapped with those of the primary cohorts. Given the small sample and absence of a prespecified inferential comparison, these data do not establish PCS specificity or demonstrate a shared post-infectious biomarker, but indicate that EV surface epitopes may also hold discrimative value on other post-viral conditions.

The longitudinal analysis showed that all six EV classification scores decreased in the 29 participants with complete model-compatible paired data. This indicates that the model-derived EV profile is time-varying. Possible explanations include longitudinal changes in circulating EV abundance, source-cell activity, or surface-epitope density; the present assay cannot distinguish among these mechanisms^32^. The raw probability decrease was descriptively larger for PS than for TSPN detection, but probabilities from separately fitted models do not share an inherently comparable scale, and standardized cross-assay comparisons did not show a significant PS-versus-TSPN difference. Importantly, score changes were not directly related to changes in fatigue, cognition, or other symptoms, and the follow-up subset may not represent the complete cohort – inherent to the fatigue component of PCS, loss of patients to follow-up and thus missing longitudinal samples remains a challenge. These findings should therefore be described as longitudinal score reduction rather than evidence that the EV signature tracks clinical course or resolution.

Beyond EV surface markers, the cargo of neuronal-enriched plasma EV preparations offers a complementary window onto the neurological burden of PCS: previousy studies showed neuronal-enriched plasma EVs from individuals recovering from COVID-19 carrying significantly elevated total tau, phosphorylated tau (p-T181), neurofilament light, neurogranin and amyloid-β^37^. This converges with non-EV-based diagnostic efforts. Plasma phosphorylated tau is emerging as an accessible marker of neurological post-acute sequelae—pTau-181 rose by approximately 59% after COVID-19 onset in essential workers who developed neurological sequelae, particularly those with persistent central symptoms^38^, and pTau-217 correlated with choroid-plexus enlargement and reduced perfusion, and these choroid-plexus abnormalities were associated with cognitive impairment in long COVID^39^. EV surface profiling, EV cargo analysis and soluble neurodegeneration markers are therefore likely to be complementary rather than competing readouts; combining an EV surface signature with neuronal-EV cargo or plasma phosphorylated tau may be especially informative for the subset of participants with objectively lower cognitive performance^33^.

Our study has several limitations. All preprocessing, regularization tuning, and adaptive CV-selected and CV-1SE panel selection were fully nested within the cross-validation folds, reducing information leakage; nevertheless, the adaptive selection procedures remain exploratory, and model development and internal evaluation were performed in the same single-centre cohort. The CV-selected and CV-1SE analyses are fold-specific feature-reduction strategies rather than fixed validated biomarker panels. The pooled clinical correlations were largely driven by between-group differences and did not remain significant within either cohort after correction or after controlling for cohort, i.e. did not correlate with severity. The bead-based assay was performed directly in double-centrifuged plasma without orthogonal EV characterization by electron microscopy, EV-marker immunoblotting, detergent lysis, or dedicated lipoprotein, haemolysis, and residual-platelet controls, as analysis in minimally processed material was the performed to reduce loss of informative EVs and exploit the immunocapturing aspect of bead-assisted flow cytometry. Consequently, bead-associated signals cannot be attributed exclusively to EVs, and residual non-vesicular or platelet-associated contributions cannot be excluded. The COVID^post^ group also included retrospective biobank cases, although the prospective-only sensitivity analysis preserved the principal classification pattern. Vaccination status was recorded at study admission, but the sequence of vaccination relative to infection was unavailable, and vaccination-subgroup effects were not estimated because few participants were unvaccinated. The non-COVID^post^ cohort was too small for formal specificity or cross-post-viral inference. Independent, preferably multicentre, validation is required before diagnostic or prognostic interpretation.

In summary, multiplex plasma EV surface profiling identified an internally cross-validated, distributed marker pattern that moderately distinguished COVID^post^ from COVID^reco^ participants. EV scores reflected pooled clinical group differences and decreased at follow-up, but the present study does not establish within-PCS severity tracking, clinical normalization, diagnostic specificity, or a validated fixed marker panel. Independent validation and prospective assessment alongside clinical phenotyping will be required to determine diagnostic or stratification value of these panels. The results support further prospective evaluation of multiparametric plasma EV surface profiling in PCS and independent validation of its diagnostic or stratification value.

## Data availability

Deidentified MESF matrices and analysis scripts will be deposited in a public repository upon publication. Clinical data are not publicly available because of participant privacy but may be made available by the corresponding authors upon reasonable request, subject to ethics and institutional approval.

## Author contributions

Conceptualization: D.K.E., J.M., H.T., and R.M.G.; Methodology: D.K.E., J.M., H.T., and R.M.G.; Investigation: D.K.E. (clinical investigation), H.R., L.T.B., S.C., D.L., S.M., and J.L.; Resources: D.K.E. and H.T. (cohort recruitment); Data curation: D.K.E., H.R., and R.M.G.; Formal analysis: D.K.E., H.R., and R.M.G.; Validation: D.K.E., H.R., J.M., H.T., and R.M.G.; Visualization: R.M.G.; Supervision: D.K.E., J.M., H.T., and R.M.G.; Project administration: D.K.E., J.M., H.T., and R.M.G.; Funding acquisition: J.M., H.T., and R.M.G.; Writing: original draft: R.M.G.; review & editing: all authors. All authors contributed to interpretation of the data, approved the submitted version.

## Acknowledgments

We want to thank Refika Aksamija, Joleene Holm, Nicole Renske, Tatiana Simak, Vera Lehmensiek, Sandra Hübsch, Dagmar Schattauer, Alice Beer, Dr. Stephanie Becker, and Martina Leis from the Laboratory of Cerebrospinal Fluid Diagnostics and Clinical Neurochemistry, the Autoimmune Laboratory, and Biobank of the Neurological Department of the University of Ulm (Germany).

## Declaration of generative AI use

During the preparation of this work, the authors used OpenAI Codex (GPT-5.6 Sol) and Anthropic Claude Opus (versions 4.8 and 5.0) for language editing and to assist with the development, execution, and verification of data-analysis code. All analytical decisions, data assignments, statistical outputs, and interpretations were independently reviewed and validated by the authors.

## References

[1] Soriano JB, Murthy S, Marshall JC, Relan P, Diaz JV. A clinical case definition of post-COVID-19 condition by a Delphi consensus. Lancet Infect Dis. 2022;22(4):e102–e107. doi:10.1016/S1473-3099(21)00703-9

[2] Davis HE, McCorkell L, Vogel JM, Topol EJ. Long COVID: major findings, mechanisms and recommendations. Nat Rev Microbiol. 2023;21(3):133–146. doi:10.1038/s41579-022-00846-2

[3] Al-Aly Z, Xie Y, Bowe B. High-dimensional characterization of post-acute sequelae of COVID-19. Nature. 2021;594(7862):259–264. doi:10.1038/s41586-021-03553-9

[4] Swank Z, Senussi Y, Manickas-Hill Z, et al. Persistent circulating severe acute respiratory syndrome coronavirus 2 spike is associated with post-acute coronavirus disease 2019 sequelae. Clin Infect Dis. 2023;76(3):e487–e490. doi:10.1093/cid/ciac722

[5] Proal AD, VanElzakker MB, Aleman S, et al. SARS-CoV-2 reservoir in post-acute sequelae of COVID-19 (PASC). Nat Immunol. 2023;24(10):1616–1627. doi:10.1038/s41590-023-01601-2

[6] Cervia-Hasler C, Brüningk SC, Hoch T, et al. Persistent complement dysregulation with signs of thromboinflammation in active Long Covid. Science. 2024;383(6680):eadg7942. doi:10.1126/science.adg7942

[7] Arron HE, Marsh BD, Kell DB, Khan MA, Jaeger BR, Pretorius E. Myalgic Encephalomyelitis/Chronic Fatigue Syndrome: the biology of a neglected disease. Front Immunol. 2024;15:1386607. doi:10.3389/fimmu.2024.1386607

[8] Yong SJ, Halim A, Halim M, et al. Inflammatory and vascular biomarkers in post-COVID-19 syndrome: a systematic review and meta-analysis of over 20 biomarkers. Rev Med Virol. 2023;33(2):e2424. doi:10.1002/rmv.2424

[9] Welsh JA, Goberdhan DCI, O’Driscoll L, et al. Minimal information for studies of extracellular vesicles (MISEV2023): from basic to advanced approaches. J Extracell Vesicles. 2024;13(2):e12404. doi:10.1002/jev2.12404

[10] Yu W, Hurley J, Roberts D, et al. Exosome-based liquid biopsies in cancer: opportunities and challenges. Ann Oncol. 2021;32(4):466–477. doi:10.1016/j.annonc.2021.01.074

[11] Li W, Liu JB, Hou LK, et al. Liquid biopsy in lung cancer: significance in diagnostics, prediction, and treatment monitoring. Mol Cancer. 2022;21(1):25. doi:10.1186/s12943-022-01505-z

[12] d’Alessandro M, Soccio P, Bergantini L, et al. Extracellular vesicle surface signatures in IPF patients: a multiplex bead-based flow cytometry approach. Cells. 2021;10(5):1045. doi:10.3390/cells10051045

[13] Barberis E, Vanella VV, Falasca M, et al. Circulating exosomes are strongly involved in SARS-CoV-2 infection. Front Mol Biosci. 2021;8:632290. doi:10.3389/fmolb.2021.632290

[14] Pesce E, Manfrini N, Cordiglieri C, et al. Exosomes recovered from the plasma of COVID-19 patients expose SARS-CoV-2 spike-derived fragments and contribute to the adaptive immune response. Front Immunol. 2022;12:785941. doi:10.3389/fimmu.2021.785941

[15] Bertrams W, Roessler FK, Bæk R, et al. Surface proteome of plasma extracellular vesicles differentiates between SARS-CoV-2 and influenza infection. Virulence. 2026;17(1):2590305. doi:10.1080/21505594.2025.2590305

[16] Seifert M, Schäfers J, Douglas FF, et al. Extracellular vesicle protein and miRNA signatures as biomarkers for post-infectious ME/CFS patients. Int J Mol Sci. 2026;27(5):2314. doi:10.3390/ijms27052314

[17] Pflueger M, Gschwandtner U. Testbatterie zur Aufmerksamkeitsprüfung (TAP) Version 1.7. Z Klin Psychol Psychother. 2003;32:155–157. doi:10.1026/0084-5345.32.2.155

[18] Smith A. Symbol Digit Modalities Test (SDMT): Manual (Revised). Los Angeles, CA: Western Psychological Services; 1982.

[19] Aschenbrenner S, Tucha O, Lange KW. Regensburger Wortflüssigkeits-Test (RWT): Handanweisung. Göttingen: Hogrefe; 2000.

[20] Elwood RW. The Wechsler Memory Scale-Revised: Psychometric characteristics and clinical application. Neuropsychol Rev. 1991;2(2):179–201. doi:10.1007/bf01109053

[21] Helmstaedter C, Lendt M, Lux S. Verbaler Lern-und Merkfähigkeitstest (VLMT): Manual. Göttingen: Beltz Test; 2001.

[22] Merten T, Blaskewitz N. Der Rey Complex Figure Test and Recognition Trial in der klinischen Praxis. Neurol Rehabil. 2008;14(4):195–202.

[23] Penner IK, Raselli C, Stöcklin M, Opwis K, Kappos L, Calabrese P. The Fatigue Scale for Motor and Cognitive Functions (FSMC): validation of a new instrument to assess multiple sclerosis-related fatigue. Mult Scler. 2009;15(12):1509–1517. doi:10.1177/1352458509348519

[24] Guilmette TJ, Sweet JJ, Hebben N, et al. American Academy of Clinical Neuropsychology consensus conference statement on uniform labeling of performance test scores. Clin Neuropsychol. 2020;34(3):437–453. doi:10.1080/13854046.2020.1722244

[25] Binder LM, Iverson GL, Brooks BL. To err is human: “abnormal” neuropsychological scores and variability are common in healthy adults. Arch Clin Neuropsychol. 2009;24(1):31–46. doi:10.1093/arclin/acn001

[26] Schretlen DJ, Munro CA, Anthony JC, Pearlson GD. Examining the range of normal intraindividual variability in neuropsychological test performance. J Int Neuropsychol Soc. 2003;9(6):864–870. doi:10.1017/S1355617703960061

[27] Tumani H, Petereit HF, Gerritzen A, et al. S1 guidelines “lumbar puncture and cerebrospinal fluid analysis” (abridged and translated version). Neurol Res Pract. 2020;2:8. doi:10.1186/s42466-020-0051-z

[28] Freedman MS, Thompson EJ, Deisenhammer F, et al. Recommended standard of cerebrospinal fluid analysis in the diagnosis of multiple sclerosis: a consensus statement. Arch Neurol. 2005;62(6):865–870. doi:10.1001/archneur.62.6.865

[29] Reiber H, Lange P. Quantification of virus-specific antibodies in cerebrospinal fluid and serum: sensitive and specific detection of antibody synthesis in brain. Clin Chem. 1991;37(7):1153–1160. doi:10.1093/clinchem/37.7.1153

[30] Reiber H. Cerebrospinal fluid - physiology, analysis and interpretation of protein patterns for diagnosis of neurological diseases. Mult Scler. 1998;4(3):99–107. doi:10.1177/135245859800400302

[31] Groß R, Reßin H, von Maltitz P, et al. Phosphatidylserine-exposing extracellular vesicles in body fluids are an innate defence against apoptotic mimicry viral pathogens. Nat Microbiol. 2024;9(4):905–921. doi:10.1038/s41564-024-01637-6

[32] Flaskamp L, Prechtl M, Scheck A, et al. Assessing extracellular vesicle turnover in vivo using highly sensitive phosphatidylserine-binding reagents. Adv Sci (Weinh). 2025;12(40):e07624. doi:10.1002/advs.202507624

[33] Balz LT, Erhart DK, Uttner I, Lulé DE, Tumani H. Evidence for a severe cognitive subgroup in a comprehensive neuropsychological Post-COVID-19 syndrome classification. Sci Rep. 2025;15(1):40368. doi:10.1038/s41598-025-25453-y

[34] Martins-Gonçalves R, Campos MM, Palhinha L, et al. Persisting platelet activation and hyperactivity in COVID-19 survivors. Circ Res. 2022;131(11):944–947. doi:10.1161/CIRCRESAHA.122.321659

[35] Gottlieb M, Spatz ES, Yu H, et al. Long COVID clinical phenotypes up to 6 months after infection identified by latent class analysis of self-reported symptoms. Open Forum Infect Dis. 2023;10(7):ofad277. doi:10.1093/ofid/ofad277

[36] Xu X, Iqbal Z, Xu L, et al. Brain-derived extracellular vesicles: potential diagnostic biomarkers for central nervous system diseases. Psychiatry Clin Neurosci. 2024;78(2):83–96. doi:10.1111/pcn.13610

[37] Sun B, Tang N, Peluso MJ, et al. Characterization and biomarker analyses of post-COVID-19 complications and neurological manifestations. Cells. 2021;10(2):386. doi:10.3390/cells10020386

[38] Yang X, Fontana A, Clouston SAP, Luft BJ. Increased phosphorylated tau (pTau-181) is associated with neurological post-acute sequelae of coronavirus disease in essential workers: a prospective cohort study before and after COVID-19 onset. EBioMedicine. 2026;123:106106. doi:10.1016/j.ebiom.2025.106106

[39] Pang H, Frontera J, Jiang L, et al. Choroid plexus alterations in long COVID and their associations with Alzheimer’s disease risks. Alzheimers Dement. 2026;22(2):e71020. doi:10.1002/alz.71020

[40] Reiber H. Flow rate of cerebrospinal fluid (CSF) - a concept common to normal blood-CSF barrier function and to dysfunction in neurological diseases. J Neurol Sci. 1994;122(2):189–203. doi:10.1016/0022-510X(94)90298-4

