## Supplement for "A Multivariable Plasma Extracellular Vesicle Surface Profile Associated with Post-COVID-19 Syndrome"

**Supplementary material**

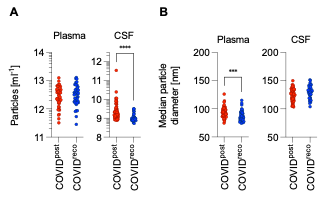

**Supplementary Figure S1.** Biophysical characterization of extracellular vesicles by nanoparticle tracking analysis. (A) Particle concentration and (B) median particle diameter were measured in EDTA plasma (left) and cerebrospinal fluid (CSF; right). Groups were compared using two-sided Mann–Whitney U tests. Plasma median particle diameter differed between groups (p=0.012), as did CSF particle concentration (p<0.001); plasma particle concentration and CSF median particle diameter were not significantly different. *p<0.05, ***p<0.001.

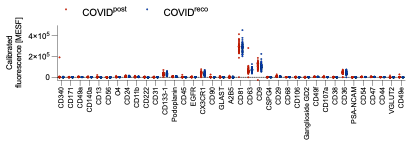

**Supplementary Figure S2.** CSF extracellular-vesicle surface profiling using a neurology-focused MACSPlex panel. CSF EVs were captured on 37 analyte-specific bead populations and detected using an APC-conjugated anti-CD9/CD63/CD81 antibody cocktail. Sample and corresponding blank fluorescence were converted separately to molecules of equivalent soluble fluorophore (MESF) using APC calibration before blank-MESF subtraction. Same-run technical duplicates were averaged; all acquired samples from the final Figure 1 population were included irrespective of bead count, and signed blank-subtracted values were retained. Red indicates COVID^post^ (nominal n=30) and blue indicates COVID^reco^ (nominal n=37); marker-specific analyzable sample sizes were n=28–30 and n=36–37, respectively, because isolated nonpositive or missing raw fluorescence values could not be log-calibrated. The panel includes markers associated with neural, glial, microglial and oligodendrocyte-lineage cells. Each dot represents one participant, and the dashed line indicates the APC LLOQ (284 MESF). No marker met the two-stage Benjamini–Krieger–Yekutieli false-discovery-rate threshold of 1% after two-sided Mann–Whitney U testing across the 37 markers.

**Table S1. Clinical symptoms of the two prospective cohorts COVID^post^ and non-COVID^post^ at initial presentation in the Post-COVID syndrome outpatient unit.**

| **Symptom** | **COVID^post^ (n=47)** | **non-COVID^post^ (n=10)** |
| --- | --- | --- |
| **Subjective cognition deficits (n), %** | **(47/47), 100%** | **(9/10), 90%** |
| **Fatigue, incl. post-exertional malaise (n), %** | **(47/47), 100%** | **(10/10), 100%** |
| **Headache (n), %** | (23/47), 48.9% | **(4/10), 40%** |
| **Dizziness (n), %** | **(8/47), 17.0%** | **(4/10), 40%** |
| **Myalgia (n), %** | **(22/47), 46.8%** | **(6/10), 60%** |
| **Arthralgia (n), %** | **(14/47), 29.8%** | **(4/10), 40%** |
| **Sleep disturbance (n), %** | (29/47), 61.7% | **(8/10), 80%** |
| **Sensory disturbance (n), %** | **(11/47), 23.4%** | **(2/10), 20%** |
| **Tremor (n), %** | (0/47), 0% | **(0/10), 0%** |
| **Chronic pain syndrome (n), %** | **(10/47), 21.3%** | **(2/10), 20%** |
| **Exertional dyspnoea (n), %** | **(12/47), 25.5%** | **(1/10), 10%** |
| **Resting dyspnoea (n), %** | **(4/47), 8.5%** | **(0/10), 0%** |
| **Postural tachycardia (HR ≥ 100 bpm) (n), %** | **(17/47), 36.2%** | **(3/10), 30%** |
| **Tinnitus (n), %** | **(4/47), 8.5%** | **(1/10), 10%** |
| **Visual disturbance (n), %** | **(4/47), 8.5%** | **(0/10), 0%** |
| **Hyposmia/Anosmia (n), %** | **(4/47), 8.5%** | **(0/10), 0%** |
| **Dysgeusia (n), %** | **(2/47), 4.3%** | **(0/10), 0%** |
| **Gait/movement disturbance (n), %** | **(2/47), 4.3%** | **(0/10), 0%** |
| **Fever/subfebrile (n), %** | **(7/47), 14.9%** | **(4/10), 40%** |
| **Altered bowel habits (diarrhea/constipation) (n), %** | **(9/47), 19.1%** | **(2/10), 20%** |
| **Abdominal pain (n), %** | **(8/47), 17.0%** | **(1/10), 10%** |
| **Nocturnal dry cough (n), %** | **(1/47), 2.1%** | **(0/10), 0%** |
| **Flu-like symptoms/recurrent URTI (n), %** | **(16/47), 34.0%** | **(4/10), 40%** |
| **Hair loss (n), %** | **(1/47), 2.1%** | **(0/10), 0%** |
| **Photophobia/phonophobia (n), %** | **(12/47), 25.5%** | **(2/10), 20%** |

**HR: heart rate, bpm: beats per minute, URTI: upper respiratory tract infections.**

**Table S2.** Composition and nested cross-validated classification performance of plasma EV surface-marker models (COVID^post^ versus COVID^reco^). TSPN- and PS-detected profiles were analyzed separately in 52 COVID^post^ and 80 COVID^reco^ participants. The All model used all 37 markers as candidate inputs. CV-selected and CV-1SE reselected marker identity and number within each outer-training set; reduced-panel sizes are medians (ranges) across the 25 outer fits and therefore do not define fixed marker signatures. Biology-based models used the fixed categories defined in Figure 2. AUC values are mean ± SD of five repeat-specific out-of-fold AUCs from five repeats of stratified fivefold nested cross-validation; SD reflects split-to-split variability. Ninety-five per cent CIs were obtained from 10,000 cohort-stratified participant-level bootstrap resamples (seed 20260826), with each participant’s five repeat-specific out-of-fold predictions resampled together. AUC was recalculated per repeat and averaged, and percentile limits are reported. These CIs are conditional on the fitted cross-validation predictions and quantify participant-sampling uncertainty rather than external-validation or full model-refitting uncertainty.

| **Model procedure** | **Detection** | **Input/selected markers** | **Panel composition** | **AUC, mean ± SD (95% CI)** |
| --- | --- | --- | --- | --- |
| **Data-driven models (nested fold-adaptive feature selection)** | | | | |
| All | TSPN | 37 candidate markers | All 37 markers; fold-specific nonzero coefficients | 0.788 ± 0.021 (0.715–0.852) |
| All | PS | 37 candidate markers | All 37 markers; fold-specific nonzero coefficients | 0.716 ± 0.026 (0.636–0.792) |
| CV-selected | TSPN | 10 (3–26) | Fold-specific; maximizes inner-CV AUC | 0.744 ± 0.021 (0.672–0.810) |
| CV-selected | PS | 23 (1–32) | Fold-specific; maximizes inner-CV AUC | 0.669 ± 0.027 (0.590–0.748) |
| CV-1SE | TSPN | 7 (2–20) | Fold-specific; smallest panel within 1 SE | 0.703 ± 0.020 (0.635–0.770) |
| CV-1SE | PS | 11 (1–22) | Fold-specific; smallest panel within 1 SE | 0.631 ± 0.034 (0.560–0.703) |
| **Biology-based models (fixed marker categories, L1-weighted)** | | | | |
| Vasculature | TSPN | 8 | CD42a, CD41b, CD62P, CD31, CD146, CD105, CD142, CD49e | 0.702 ± 0.022 (0.614–0.782) |
|  | PS |  |  | 0.615 ± 0.023 (0.520–0.710) |
| Stem | TSPN | 4 | CD133-1, ROR1, MCSP, SSEA-4 | 0.661 ± 0.004 (0.569–0.749) |
|  | PS |  |  | 0.630 ± 0.024 (0.538–0.720) |
| Immunity | TSPN | 14 | CD3, CD4, CD8, CD19, CD20, CD1c, CD209, HLA-DR/DP/DQ, HLA-ABC, CD86, CD56, CD69, CD25, CD45 | 0.576 ± 0.024 (0.489–0.659) |
|  | PS |  |  | 0.624 ± 0.023 (0.536–0.710) |
| Other | TSPN | 8 | CD2, CD14, CD11c, CD40, CD29, CD44, CD326, CD24 | 0.570 ± 0.016 (0.483–0.658) |
|  | PS |  |  | 0.588 ± 0.036 (0.502–0.675) |
